# Identifying Circulating Protein Mediators in the Link Between Smoking and Abdominal Aortic Aneurysm: An Integrated Analysis of Human Proteomic and Genomic Data

**DOI:** 10.1101/2025.02.27.25322973

**Authors:** Shuai Yuan, Samuel Khodursky, Jiawei Geng, Pranav Sharma, Joshua M. Spin, Philip Tsao, Michael G. Levin, Scott M. Damrauer

## Abstract

**Background:** Smoking is a well-established risk factor for abdominal aortic aneurysm (AAA). However, the molecular pathways underlying this relationship remain poorly understood. This study aimed to identify circulating protein mediators that may explain the association between smoking and AAA.

**Methods:** We conducted a network Mendelian randomization (MR) study utilizing summary-level data from the largest available genome-wide association studies. Our primary smoking exposure was the lifetime smoking index, with smoking initiation and cigarettes per day included as supplementary traits. The AAA dataset comprised 39,221 cases and 1,086,107 controls. Protein data were sourced from two large cohorts: UKB-PPP, where proteins were measured using the Olink platform in 54,219 individuals, and deCODE, where proteins were measured using the SomaScan platform in 35,559 individuals. Two-sample MR was employed to estimate the association between smoking and AAA (β_total_) and between smoking and circulating protein levels (β_1_). Summary data-based MR was then used to assess the association between smoking-related proteins and AAA risk (β_2_). Mediation pathways were identified based on the directionality of effect estimates, and the corresponding mediation effects were quantified.

**Results:** Genetically predicted smoking traits were consistently associated with an increased risk of AAA. The lifetime smoking index was associated with the levels of 543 out of 5,764 unique circulating proteins, with 470 of these associations replicated in supplementary analyses using additional smoking traits and protein sources. Among the smoking-related proteins, genetically predicted levels of 22 were associated with AAA risk. Eight mediation pathways were identified accounting for 42.7% of the total smoking-AAA association and with mediation effects >4% for ADAMTS15, IL1RN, MMP12, PGF, PCSK9, and UXS1.

**Conclusion:** This study identified numerous circulating proteins potentially causally linked to smoking, and eight of these proteins were found to mediate the association between smoking and AAA risk.

## Introduction

Abdominal aortic aneurysm (AAA) is a potentially life-threatening condition characterized by the abnormal enlargement of the abdominal aorta, which, if left untreated, can lead to rupture. AAA is typically asymptomatic until rupture occurs, a critical event that is often fatal without timely surgical intervention. In 2017, AAA was responsible for 167,200 deaths and 3 million disability-adjusted life years worldwide, with the majority of cases occurring in individuals over the age of 50.^1,2^

Smoking is one of the most significant risk factors for AAA, contributing to both the development and progression of the disease.^3–5^ The mechanisms underlying the smoking-AAA link appear to involve long-term alterations in vascular smooth muscle cells and inflammatory cell function.^6^ However, the detailed molecular mechanisms remain unclear due to conflicting findings and limited evidence from clinical studies in humans. For instance, increased matrix metalloproteinases (MMP) expression following cigarette smoke exposure is thought to contribute to AAA development,^7^ yet some studies in mice suggest that smoking’s impact may be independent of elastolytic enzymes.^8,9^ Given that circulating proteins are potential therapeutic targets, understanding the protein pathways linking smoking to AAA could help identify new strategies for preventing or treating AAA, especially in smokers.

With the increasing availability of large-scale genomics and proteomics data, a genetics-based approach, particularly Mendelian randomization (MR) linking proteins to disease has become a widely adopted method for therapeutic discovery,^10–12^ demonstrating high success rates^13,14^. In this study, we integrated human proteomic and genomic data from large-scale cohorts to investigate the protein pathways underlying the elevated risk of AAA associated with cigarette smoking, using a two-stage network MR framework.

## Methods

### Study design

Figure 1 provides an overview of the study design and data sources utilized. Detailed information on used data sources is given in **Table S1**. Our network MR analysis involved three key steps.^15^ First, we estimated the total effect of genetically predicted smoking on the genetic liability to AAA. Next, we examined associations between genetically predicted smoking and levels of 5,764 circulating proteins. Finally, we investigated the associations between genetically predicted levels of smoking-related proteins and the risk of AAA. Based on these analyses, we constructed pathways and quantified the mediation effects, offering insights into the molecular mechanisms linking smoking to AAA.

**Figure 1.**
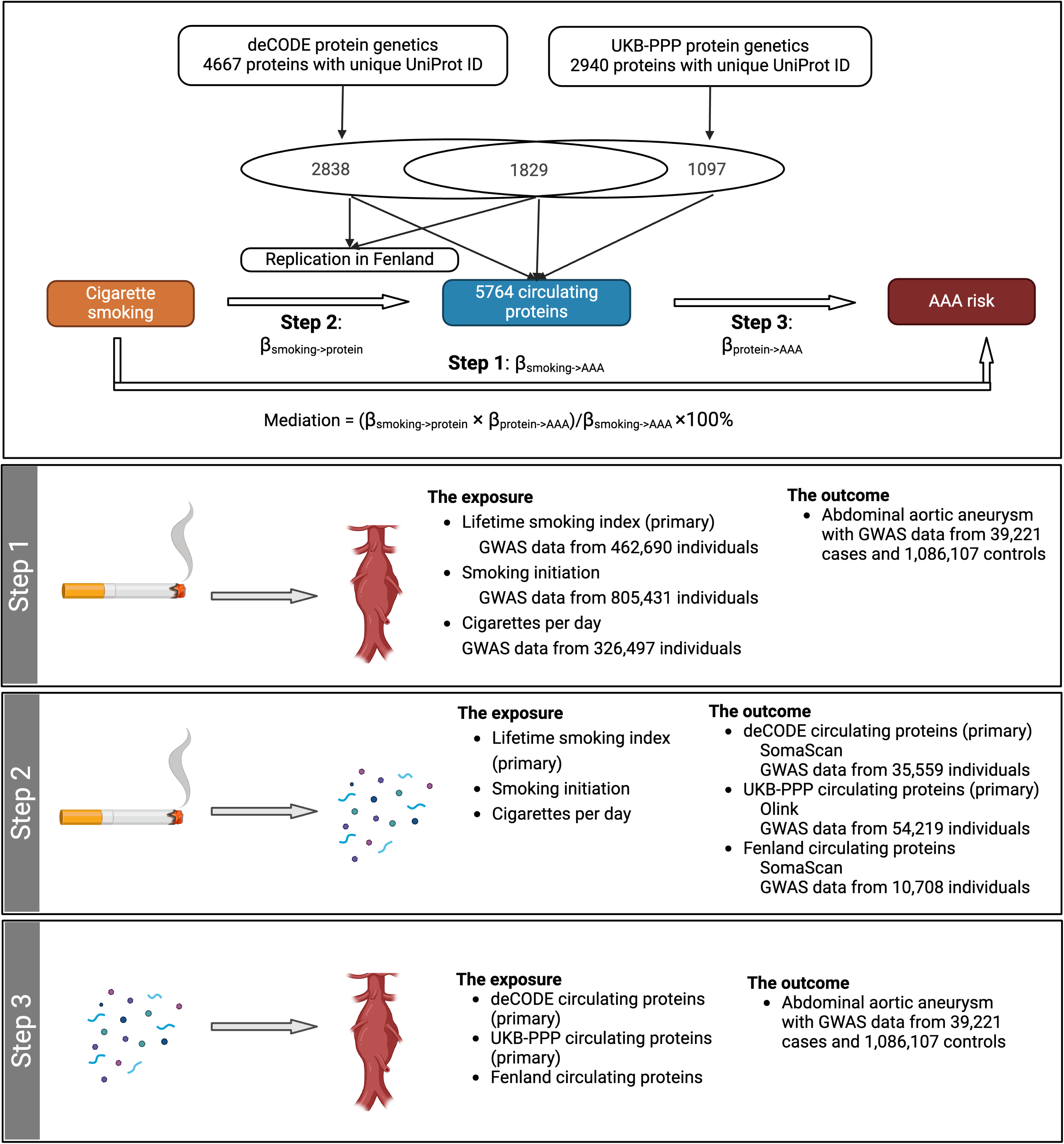
Study design overview. AAA, abdominal aortic aneurysm; GWAS, genome-wide association study; UKB-PPP, UK Biobank Pharma Proteomics Project.

### Data sources for smoking traits

We used the lifetime smoking index as the primary measure of smoking exposure, and smoking initiation and cigarettes per day as secondary measures. The lifetime smoking index is a comprehensive measure of an individual’s overall smoking exposure, accounting for both the intensity and duration of smoking habits.^16^ Genome-wide association study (GWAS) of lifetime smoking index included 462,690 individuals, yielding 126 SNPs as instrumental variables based on the criteria of *P* < 5×10^-8^ and linkage disequilibrium *r^2^* <0.001.^16^ For smoking initiation and cigarettes per day, GWAS data including 805,431 and 326,497 individuals from GSCAN (GWAS & Sequencing Consortium of Alcohol and Nicotine use), respectively, identified 248 and 53 SNPs under identical criteria.^17^ Detailed information on these genetic variants is presented in **Table S2**.

### Data sources for AAA

GWAS summary statistics for AAA were sourced from the AAAgen consortium, a large GWAS meta-analysis involving 39,221 cases and 1,086,107 controls.^18^ AAA was defined using International Classification of Disease codes, with diagnostic data collected from hospital records or through imaging studies, depending on the specific studies included in the consortium.^18^

### Data sources for circulating proteins

GWAS data on circulating proteins were primarily obtained from two key sources: the UK Biobank Pharma Proteomics Project (UKB-PPP)^19^ and deCODE^20^. UKB-PPP analyzed 2,940 unique circulating proteins in 54,219 individuals using the Olink platform,^19^ while deCODE examined over 4,970 plasma proteins in 35,559 Icelanders using the SomaScan platform^20^. For proteins with identical UniProt IDs in deCODE, one protein was retained based on stronger associations with the lifetime smoking index although the associations for these paired proteins were consistent across 80.5% of cases. Overlapping proteins between the two datasets were sourced from UKB-PPP due to its larger sample size. The final analysis included 5,764 proteins. We divided these proteins into three distinct protein sets: overlapping proteins between UKB-PPP and deCODE (set 1), proteins unique to UKB-PPP (set 2), and proteins unique to deCODE (set 3).

This division was made to accommodate the varying sensitivity analyses applied to each dataset. In addition, the Fenland study served as an independent replication dataset for set 1 and 3. In this study, 3,892 proteins were measured in 10,708 individuals using the SomaScan platform.^21^

### Statistical analysis

F statistic was calculated to measure the strength of the instrumental variables for smoking traits and proteins and found to be >10, indicating limited weak instrument bias caused by sample overlap.^22^ Data were harmonized based on both effect and non-effect alleles. Genetic variants with allele mismatched were removed from the analysis.

The inverse variance weighted method under the multiplicative random effects were used as the primary analysis to estimate the smoking-AAA (β_total_) and smoking-protein (β_1_) associations and supplemented by the weighted median,^23^ weighted mode,^24^ MR-Egger^25^, and MR-PRESSO^26^. We conducted additional sensitivity analyses using various smoking-related traits and different sources of circulating proteins. An association was deemed replicated if it reached nominal significance in any of these analyses. The Cochran’s Q test was used to measure the heterogeneity of the association, and the MR-Egger intercept test was used to examine the horizontal pleiotropy. To investigate the biological pathways associated with smoking-related circulating proteins, we utilized the Enrichr online platform^27^ to perform pathway enrichment analysis based on Gene Ontology (GO) databases, including GO Biological Process, Cellular Component, and Molecular Function.

The associations between circulating proteins and AAA (β_2_) were estimated using the summary-based Mendelian Randomization (SMR) method.^28^ The analysis was based on genetic variants in the *cis* gene region for each protein. The HEIDI (heterogeneity in dependent instruments) analysis in SMR is a statistical test used to distinguish pleiotropy from linkage by assessing heterogeneity in SNP effects. To differentiate between pleiotropy and linkage, the HEIDI method was applied.^28^ This approach tests whether multiple SNPs in LD exhibit consistent effects, rejecting the causal hypothesis if significant heterogeneity is detected. To examine whether the protein and AAA share the same causal variant, we performed Bayesian colocalization analysis^29^ and Sum of Single Effects (SuSiE) regression for proteins associated with AAA after multiple testing correction.^30^ The association with posterior probabilities for hypothesis 4 (PPH4) greater than 0.7 was considered strong evidence of colocalization. We also visualized the linkage disequilibrium between SNPs within the ±1 Mb cis-coding regions for proteins and AAA using locus plots.

The mediation pathway was defined by aligning the direction of the total effect (βtotal) with the effect mediated through the protein (β1 × β2). The proportion of the effect mediated by a protein was quantified by multiplying the estimated impact of the lifetime smoking index on protein levels with the estimated impact of protein levels on AAA. To accurately estimate the standard error (SE) associated with the mediation effect, the propagation of error method, also known as the delta method, was employed.^31^ The networks between proteins involved in the pathways were visualized using the STRING database.^32^ The false discovery rate (FDR) method was used as the multiple testing correction strategy for all analyses (*P* < 0.05).

## Results

### Smoking and AAA

Genetically predicted lifetime smoking index was positively associated with the risk of AAA. Per standard deviation (SD) increase in genetically predicted lifetime smoking index, the odds ratio (OR) of AAA was 3.08 (95% CI 2.56-3.69). The associations remained consistent in the sensitivity analyses and the analysis using smoking initiation and cigarettes per day as the exposures (Figure 2). Significant horizontal pleiotropy was detected in the analyses of lifetime smoking index and smoking initiation (*P* for MR-Egger intercept test <0.05); however, the association persisted in the MR-Egger regression after correction for horizontal pleiotropy as well as in the MR-PRESSO analysis after removal of outlying genetic variants which displayed potential horizontal pleiotropy (Figure 2).

**Figure 2.**
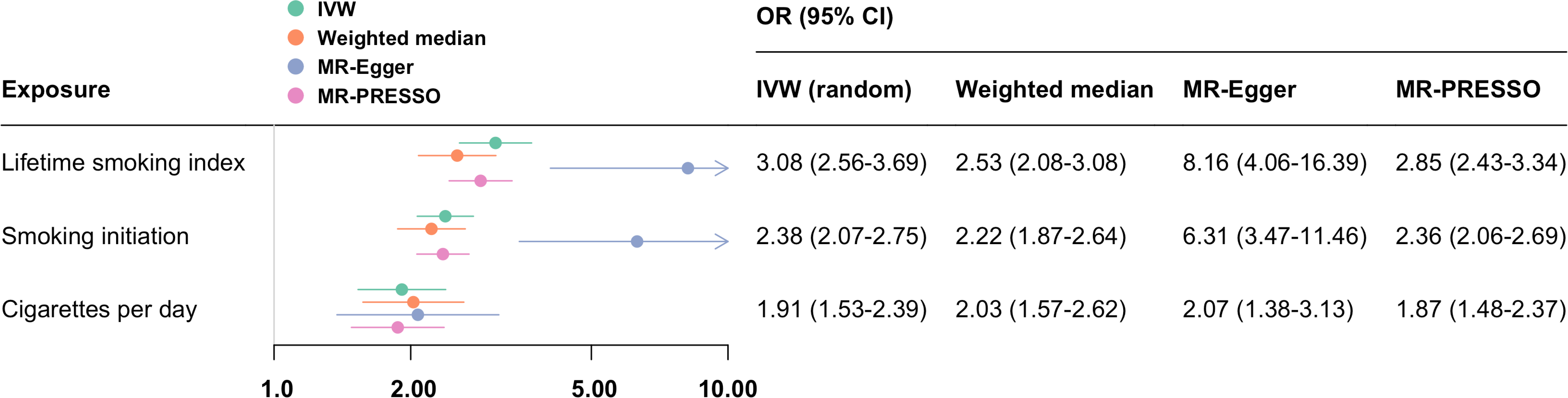
Genetically predicted smoking traits associated with genetic liability to abdominal aortic aneurysm. CI, confidence interval; IVW, inverse variance weighted; MR-PRESSO, Mendelian Randomization Pleiotropy RESidual Sum and Outlier; OR, odds ratio.

### Smoking and circulating proteins

In set 1, which included overlapping proteins between UKB-PPP and deCODE, genetically predicted lifetime smoking index was associated with levels of 412 circulating proteins after FDR correction (Figure 3a**, Table S3**). Of these, 377 associations were successfully replicated in the sensitivity analyses (**Table S4**). In set 2, comprising unique proteins from UKB-PPP, genetically predicted lifetime smoking index was associated with levels of 101 circulating proteins after FDR correction (Figure 3b**, Table S5**), with 64 of these associations replicated across sensitivity analyses using alternative smoking traits as exposures (**Table S6**). In set 3, which includes unique proteins from deCODE, genetically predicted lifetime smoking index was associated with levels of 30 circulating proteins after FDR correction (Figure 3c**, Table S7**), and 29 of these associations were successfully replicated in the sensitivity analyses (**Table S8**). Combining the significant associations across all three sets, genetically predicted lifetime smoking index was linked to levels of 470 (441 from UKB-PPP and 29 from deCODE) circulating proteins. Per standard deviation (SD) increase in genetically predicted lifetime smoking index, the change of protein levels ranged from −0.50 (95% CI −0.78, −0.23) SD for NCAM1 to 0.52 (95% CI 0.42-0.63) SD for CXCL17. The results of the supplementary analyses of these associations are presented in **Table S3-S8**. A total of 490 biological process, 46 cellular component, and 60 molecular function pathways were enriched based on the 470 smoking-associated proteins (**Table S9**). The top pathways were linked to inflammation in biological process, the collagen-containing extracellular matrix in cellular component, and endopeptidase inhibitor activity in molecular function.

**Figure 3.**
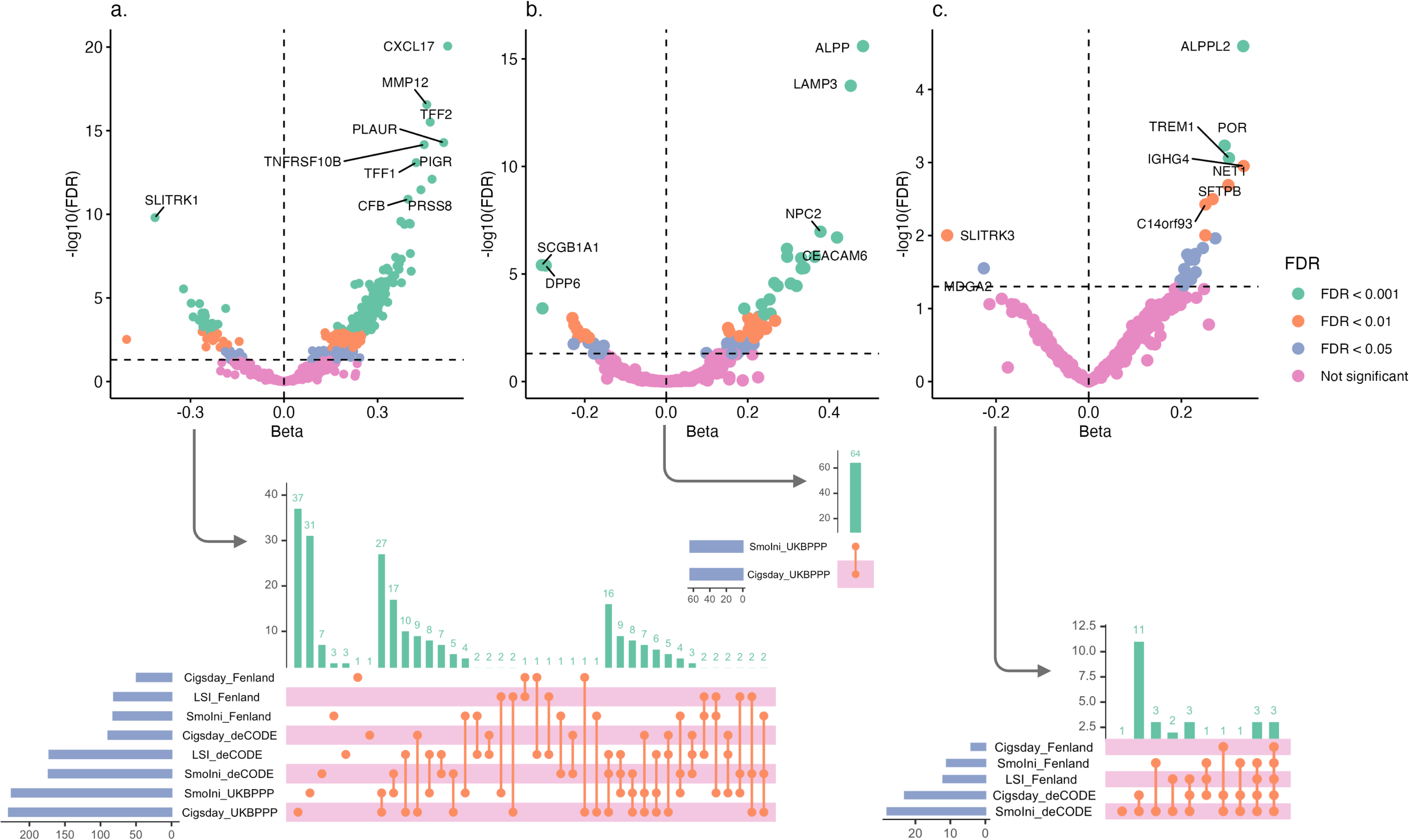
Genetically predicted smoking traits associated with genetic predicted levels of circulating proteins. FDR, false discovery rate. Panel a. the associations for overlapped proteins between UKB-PPP and deCODE (the associations were based on data from UKB-PPP). Panel b. the associations for unique proteins in UKB-PPP. Panel c. the associations for unique proteins in deCODE.

### Smoking-related proteins and AAA

Given the sample size difference between UKB-PPP and deCODE, we assessed the associations between smoking-related proteins and risk of AAA separately. Among 441 smoking-related proteins from UKB-PPP, 393 proteins with genetic instruments were included (**Table S10**). Genetically predicted levels of 22 proteins were associated with AAA (Figure 4a **and Table S10**).

**Figure 4.**
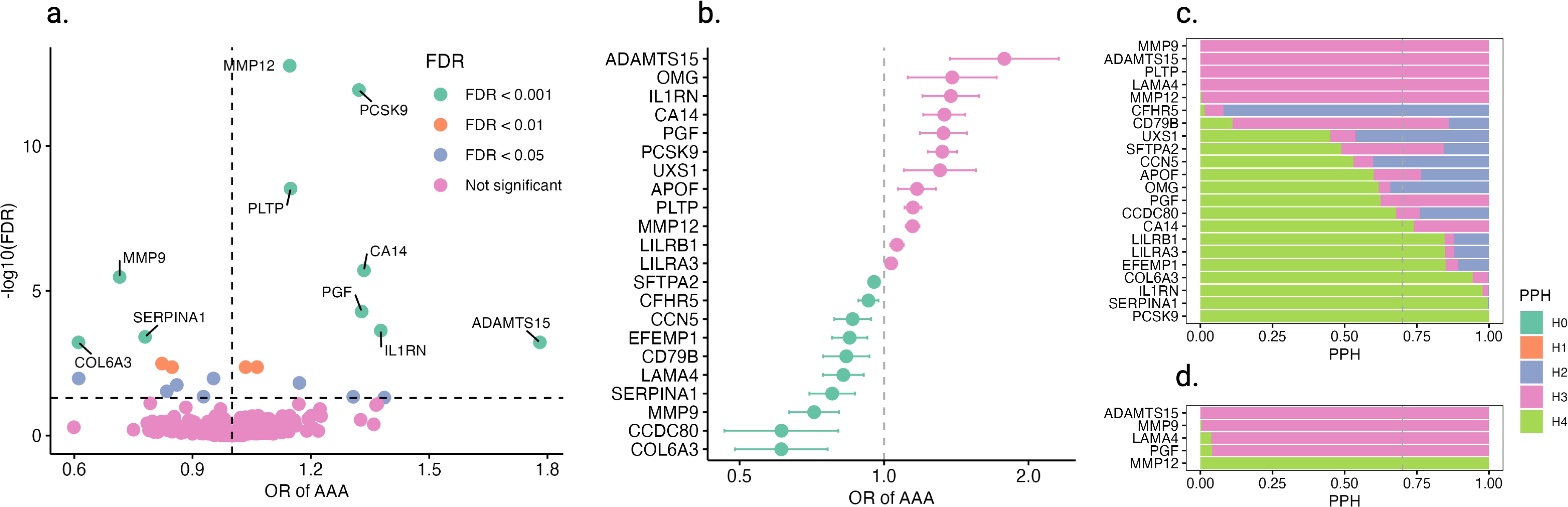
Genetically predicted smoking-related circulating proteins associated with genetic liability to abdominal aortic aneurysm. FDR, false discovery rate; OR, odds ratio; PPH, posterior probabilities for hypothesis. Panel a. volcano plot of the associations between 393 smoking-related circulating proteins and AAA risk. Panel b. forest plot of 22 associations with FDR <0.05. Panel c. results of traditional colocalization analysis. Panel d. results of SuSiE colocalization analysis.

The odds ratios (OR) for AAA ranged from 0.61 (95% CI 0.49-0.76) per standard deviation increase in genetically predicted levels of COL6A3 to 1.78 (95% CI 1.37-2.31) per standard deviation increase in genetically predicted levels of ADAMTS15 (Figure 4b). We observed significant HEIDI results for PLTP, MMP9, and PGF (*P* < 0.05), indicating potential pleiotropy for these associations (**Table S10**). Nine protein-AAA associations with PP4 >0.7 were regarded strong evidence of colocalization in traditional colocalization (Figure 4c **and Table S11**) or SuSiE colocalization analysis (Figure 4d **and Table S12**), including PCSK9, SERPINA1, IL1RN, COL6A3, EFEMP1, LILRA3, LILRB1, CA14, and MMP12. These colocalized associations were also observed in locus plots (**Figure S1**). Regarding the analysis in deCODE, 16 out of 29 proteins had genetic instruments. Genetically predicted levels of none of the 16 proteins were associated with the risk of AAA in deCODE (**Table S13**).

### Pathway network and mediation

Among the 22 smoking-related proteins associated with AAA, 8 pathways were identified where the direction of the total effect aligned with the effect mediated through the protein (Figure 5). Six proteins—ADAMTS15, IL1RN, MMP12, PGF, PCSK9, and UXS1—each mediated more than 4% of the association between the lifetime smoking index and AAA (Figure 5). These proteins exhibited limited interaction networks, except for a potential co-expression between IL1RN and MMP12 (**Figure S2**). In total, 8 proteins mediated 42.7% of the smoking-AAA association.

**Figure 5.**
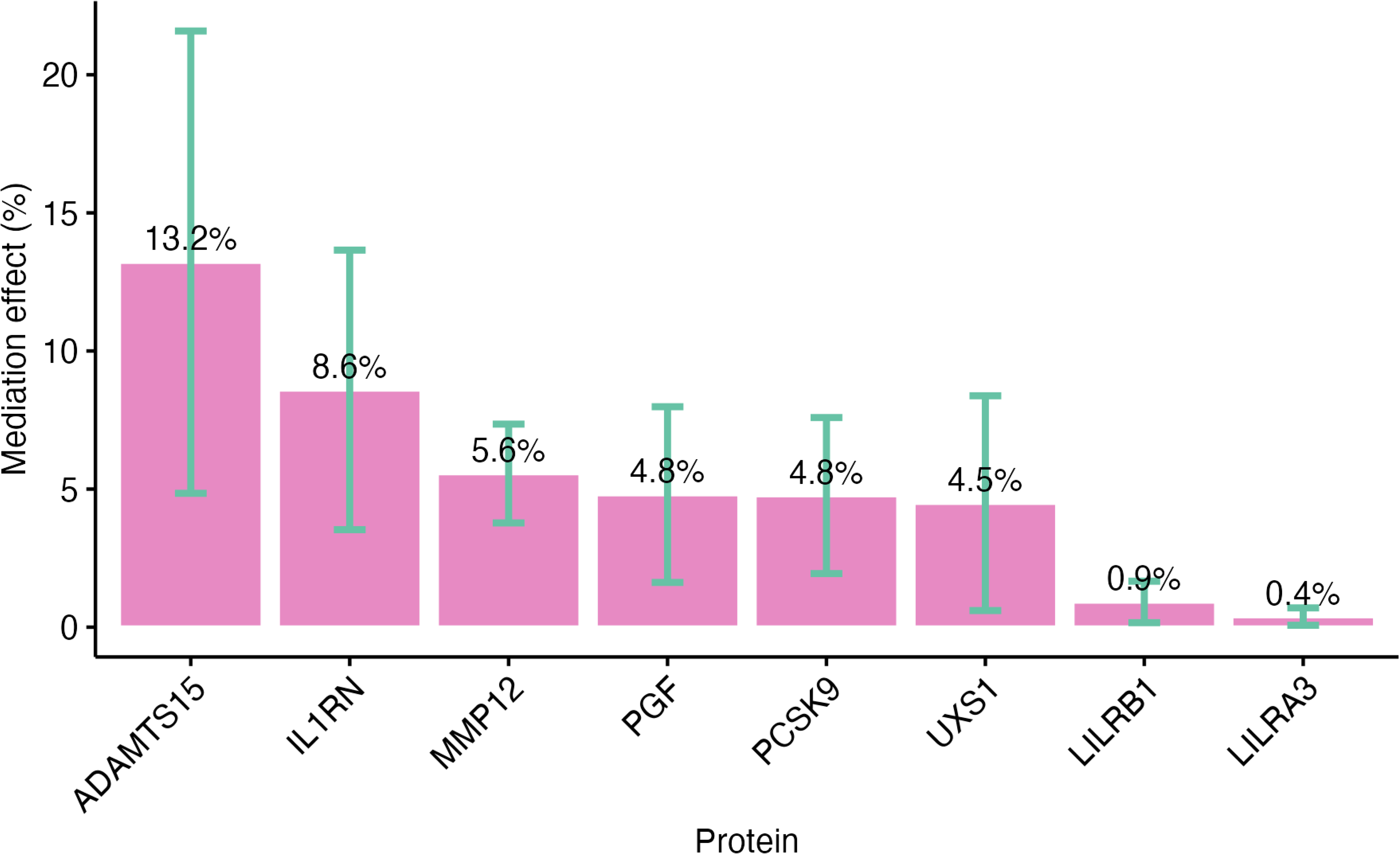
Proportion mediated by circulating proteins in the association between lifetime smoking index and risk of abdominal aortic aneurysm.

## Discussion

This large-scale network MR study investigated the associations between cigarette smoking, 5,764 circulating proteins, and the risk of AAA using proteomic and genomic data. We found that genetically predicted smoking was associated with an increased risk of AAA and levels of 470 proteins. Of these smoking-related proteins, genetically predicted levels of 22 were linked to AAA risk, with nine showing strong evidence of colocalization. Our analysis identified 8 protein pathways mediating the relationship between smoking and AAA, accounting for 42.7% of the total smoking-AAA association. This study provides a comprehensive description of the causal protein profiles influenced by cigarette smoking and highlights the protein pathways that connect smoking with AAA. Given that smoking is a major risk factor for AAA, our findings suggest potential therapeutic targets for this high-risk population.

The impact of cigarette smoking on circulating proteins has been explored in a few population-based studies. In a study involving two Swedish cohorts, current smoking was associated with 30 blood proteins, including elevated levels of MMP-12 and growth/differentiation factor 15 (GDF-15).^33^ Similarly, the Framingham Heart Study, which included 897 middle-aged adults, identified significant associations between current smoking and 53 proteins, as well as 49 proteins linked to the number of cigarette packs smoked.^34^ In the current study, we systematically examined the associations between three distinct smoking traits and 5,764 circulating proteins using a causal inference framework. Our findings confirmed many previously discovered associations and uncovered several unreported links. Among 470 smoking-associated proteins, we further uncovered potential biological, cellular, and molecular pathways, including inflammation, the collagen-containing extracellular matrix, and endopeptidase inhibitor activity. This investigation not only enhances our understanding of smoking’s effect on individual blood proteins, but also captures its broader health impact, possibly illuminating the overall pathways from smoking to a wide range of health disorders.

Mediation analysis detected eight proteins linking smoking to AAA, including ADAMTS15, IL1RN, MMP12, PGF, PCSK9, and UXS1 with mediation proportion above 4% and LILRB1 and LILRA3 with limited mediation effects. Eight proteins in total mediated 42.7% of the smoking-AAA association.

Two key protein mediators, ADAMTS15 (ADAM metallopeptidase with thrombospondin type 1 motif 15) and MMP12 (matrix metalloproteinase-12), in the smoking-AAA pathway are part of the metalloproteinase family,^35^ enzymes that are crucial for extracellular matrix remodeling. Increased MMP12 expression has been consistently observed in smokers.^33,35^ However, the impact of smoking on ADAMTS15 expression has been minimally characterized. The overexpression of these two matrix-degrading enzymes may weaken the aortic wall, contributing to aneurysm formation and progression. In keeping with this, genetic studies have also linked the genes encoding the two proteins to AAA development^18^ and aortic diameter expansion^36^, and ADAMTS15 has been linked to intracranial aneurysms^37^. The role of MMP12 in AAA has been also observed in mouse experiments.^38,39^ This study is the first to quantitatively assess the mediating roles of ADAMTS15 and MMP12 in the association between smoking and AAA. However, further research is needed to determine whether targeting these proteins with drugs could aid in the prevention or treatment of AAA among smokers. Currently, no specific drugs directly target ADAMTS15, and while broad-spectrum MMP inhibitors, such as Marimastat and Batimastat, have been developed, none are approved for routine use due to side effects from inhibiting multiple MMPs. Doxycycline is an antibiotic that has MMP inhibition and prevents AAA in mouse models; however it did not significantly reduce growth of AAA in randomized clinical trials.^40^ Notably a recent study demonstrated that MMP12 inhibition using RXP470.1 demonstrated benefit in a murine AAA model,^41^ but conflicting animal data have also been published.^42^

The role of IL1RN (interleukin-1 receptor antagonist protein) and PGF (placenta growth factor) as mediators between smoking and AAA likely centers on their involvement in vascular inflammation. Smoking has been shown to influence the levels of both IL1RN^43^ and PGF^44^.

Studies suggest that blocking the IL-1 receptor, which inhibits both IL-1α and IL-1β pathways, may be a promising therapeutic target for AAA.^45^ While this seems contrary to our findings, it aligns when considering that IL1RN levels typically rise in parallel with IL-1α and IL-1β due to feedback mechanisms, especially in generally healthy populations not receiving IL-1 pathway inhibitors. This supports the idea that drugs targeting interleukin-1 signaling to reduce vascular inflammation could be a potential treatment for AAA.^46^

Although no direct evidence links PGF to AAA, it is a member of the vascular endothelial growth factor (VEGF) family and promotes angiogenesis and recruits inflammatory cells, which contribute to vascular inflammation and matrix degradation.^47^ Elevated levels of PGF, driven by smoking-induced inflammation, may increase collagen-degrading enzyme activity and plaque instability, playing a significant role in AAA progression.^48^ Thus, aflibercept, which acts as a decoy receptor for both VEGF and PGF, may represent a potential therapeutic option for AAA; however, the side effects should be assessed.

PCSK9 (proprotein convertase subtilisin/kexin type 9) plays a key role in cholesterol metabolism by promoting the degradation of LDL receptors, thereby increasing circulating LDL cholesterol levels. Previous studies have highlighted the central role of non-high-density lipoprotein (non-HDL) cholesterol in AAA,^18^ and SNPs in the gene encoding PCSK9 have been linked to AAA^49^. A GWAS meta-analysis, combined with protein-wide MR studies and mouse models, further identified the potential for repurposing PCSK9 inhibitors for AAA prevention.^18^ Notably, cigarette smoking has been shown to stimulate PCSK9 production.^50^ Our mediation analysis suggests that PCSK9 may mediate the association between smoking and AAA.^18^ These findings collectively support the potential of PCSK9 inhibitors as promising therapeutic targets,^51^ particularly among smokers at risk of AAA UXS1 (UDP-glucuronic acid decarboxylase 1) is a relatively understudied protein, primarily recognized for its role in cancer.^52^ To date, minimal evidence has linked UXS1 to AAA.

Further research is needed to explore how this protein may mediate the association between smoking and AAA. Notably however, published data examining aortic transcriptional profiling in mouse models of AAA have identified significant differential regulation of *UXS1* in aneurysmal tissue when compared with controls (which is also the case for *MMP12*, *IL1RN*, *PSCK9* and Pgf).^18,42,53^

This comprehensive study assesses the causal effects of cigarette smoking on circulating proteins and explores the protein pathways linking smoking to AAA, leveraging data from large-scale biobanks and studies. However, there are several limitations. First, the study was based on European populations, which may confine the generalizability of these findings to non-Europeans. Second, although we performed a series of sensitivity and supplementary analyses to ensure the consistency of our results and minimize the influence of horizontal pleiotropy, we cannot fully exclude the possibility that some associations are non-causal and driven by pleiotropy, particularly for the complex behavioral traits related to smoking. Additional studies using diverse designs and datasets are needed to validate and triangulate our findings. Lastly, we used protein measurements from different proteomic platforms, which could lead to discrepancies in associations. However, we found that the results were generally consistent between analyses using SomaScan and Olink data.

In summary, this study suggests that cigarette smoking may have a broad causal impact on the circulating protein profile. Network MR analysis identifies ADAMTS15, IL1RN, MMP12, PGF, PCSK9, and UXS1 as key proteins mediating the increased risk of AAA among smokers. These findings pave the way for future research to evaluate the efficacy of targeting these proteins in preventing AAA and slowing abdominal aortic growth.

## Data Availability

All data produced in the present study are available upon reasonable request to the authors.

## Acknowledgments

We want to acknowledge the participants and investigators of the included GWASs.

## Authors’ contributions

S.Y. and S.M.D. conceived and designed the study. S.Y. and J.G. contributed to data collection and undertook the statistical analyses. S.Y. created the data visualizations and authored the initial draft of the manuscript. S.Y., S.K., J.G., P.S., J.M.S., P.T., M.G.L., and S.M.D. contributed to data interpretation, offered significant intellectual insights to the manuscript, and approved its final version.

## Data and code sharing statement

The analyses were based on publicly available data and software.

## Supplementary materials

Figure S1. Locus plots of 22 proteins and AAA.

Figure S2. Interaction networks between protein mediators of the smoking-AAA link.

Table S1. Data sources.

Table S2. Genetic instrumental variables for smoking traits.

Table S3. Associations between genetically predicted lifetime smoking index and 1829 proteins overlapped between UKB-PPP and deCODE.

Table S4. Associations between genetically predicted different smoking traits and 412 lifetime smoking index-related proteins overlapped between UKBPPP and deCODE.

Table S5. Associations between genetically predicted lifetime smoking index and 1097 proteins unique in UKB-PPP.

Table S6. Associations between genetically predicted different smoking traits and 101 lifetime smoking index-related proteins unique in UKBPPP.

Table S7. Associations between genetically predicted lifetime smoking index and 2838 proteins unique in deCODE.

Table S8. Associations between genetically predicted different smoking traits and 30 lifetime smoking index-related proteins unique in deCODE.

Table S9. GO pathways enriched based on 470 smoking-related proteins

Table S10. Associations between genetically predicted levels of smpking-related proteins in UKB-PPP and AAA risk.

Table S11. Traditional colocalization analysis of 22 protein-AAA associations.

Table S12. SuSiE colocalization analysis of 22 protein-AAA associations.

Table S13. Associations between genetically predicted levels of smpking-related proteins in deCODE and AAA risk.

